# Maternal Outcomes of Postpartum Hemorrhage After Vaginal Delivery in Lusaka, Zambia: A Hospital-Level Comparison

**DOI:** 10.64898/2026.01.14.26344145

**Authors:** Astridah K.Y. Maseka, Choongo Mulungu

## Abstract

**Introduction:** Postpartum hemorrhage is the leading contributor to maternal morbidity and mortality worldwide, with the burden greatest in low-resource settings. In Zambia, maternal mortality remains above national and global targets, and PPH is a major contributor. While most studies emphasize predictors and prevalence, limited evidence exists on maternal outcomes across different levels of care. This study aimed to examine maternal outcomes of Postpartum hemorrhage following vaginal delivery in first-level and tertiary hospitals in Lusaka.

**Methods:** We employed a quantitative, case-control study cross-sectionally on 318 women who delivered vaginally at first-level and tertiary hospitals in Lusaka, Zambia. Data were collected using structured questionnaires and analyzed with descriptive statistics, correlations, and regression. Cases and controls were comparable across baseline variables

**Results:** Among Postpartum hemorrhage cases, 56.8% required blood transfusion, 11% developed hemorrhagic shock, 3.8% underwent hysterectomy, and 3.8% died. Adverse outcomes occurred exclusively among cases. Accessibility and provider competence were negatively correlated with complications, while regression analysis identified uterine atony (OR = 2.8, 95% CI: 1.4–5.6), referral delays >2 hours (OR = 3.5, 95% CI: 1.7–7.2), and lack of blood availability (OR = 4.2, 95% CI: 2.0–8.9) as the strongest predictors of the mortality outcome.

**Conclusion:** Postpartum hemorrhage imposes a severe burden on first-level hospitals, with blood transfusion demand, referral delays, and systemic gaps driving poor outcomes. Strengthening blood bank infrastructure, improving referral pathways, and continuous provider training are critical to reducing preventable maternal deaths.

## I. INTRODUCTION

Postpartum hemorrhage (PPH) remains the leading direct cause of maternal mortality worldwide, accounting for approximately one-quarter of maternal deaths annually.^[1]^ Defined as loss of blood equal to 500 mls or more within 24 hours after vaginal delivery or 1000 mls at caesarean section,^[2]^ PPH is an obstetric emergency that requires immediate recognition and attention, as delay in treating it is an important cause of maternal mortality. ^[3,4]^ Despite advances in obstetric care, women continue to die from preventable causes of hemorrhage, particularly in low-resource settings where health system constraints exacerbate the burden.^[5]^ Say et al.,^[6]^ noted that worldwide, postpartum hemorrhage accounted for 8% of maternal deaths in developed regions of the world and 20% of maternal deaths in developing regions. Later estimates suggested that the incidence of PPH following vaginal delivery ranged between 6% and 10%, with significant variation across regions depending on access to skilled birth attendants, timely interventions, and health infrastructure.^[7]^

In Sub-Saharan Africa (SSA), PPH contributes disproportionately to maternal mortality, with studies reporting high incidence and poor outcomes due to delays in recognition, inadequate blood transfusion services, and limited surgical capacity.^[8,9]^ Zambia reflects this regional challenge, where maternal mortality remained unacceptably high despite national and global commitments. The Zambia National Health Strategic Plan (2017–2021) set a target to reduce the maternal mortality ratio (MMR) from 398 deaths per 100,000 live births in 2014 to 162 per 100,000 by 2021, while aligning with the Sustainable Development Goal (SDG) 3.1 target of fewer than 70 deaths per 100,000 live births by 2030. However, reviews after the 2021 target showed that Zambia’s MMR remained above 300 per 100,000 live births, with PPH as a major contributor.^[10,11]^

Studies in Zambia and similar low-resource contexts highlighted systemic barriers that worsen PPH outcomes. These include shortages of trained midwives and anesthetists, weak referral systems, limited blood bank capacity, and inadequate infrastructure at first-level hospitals.^[12,13]^ Accessibility gaps further compounded the problem, as many women delivered in facilities lacking emergency obstetric care, leading to delays in life-saving interventions. ^[14]^ Even where active management of the third stage of labor was practiced, inconsistent adherence to guidelines and lack of resources undermined effectiveness.^[15]^

Efforts to strengthen maternal health systems in Zambia, such as training programs and guideline dissemination, have not sufficiently reduced mortality from PPH.^[16,17]^ The persistence of high maternal deaths underscores the need for context-specific evidence on outcomes, particularly comparing primary healthcare-level hospitals with tertiary hospitals. While global literature has advanced understanding of predictors and risk factors, ^[16,18,19]^ there remains a dearth of empirical studies in Zambia focusing on maternal outcomes following PPH. Addressing this gap is critical for informing policy, resource allocation, and clinical practice to accelerate progress toward national and global maternal mortality reduction targets.

This study therefore examined maternal outcomes of postpartum hemorrhage after vaginal delivery in first-level and tertiary hospitals in Lusaka, Zambia. By analyzing outcomes across different levels of care, the research aimed to provide evidence to guide interventions that improve accessibility, provider competence, and system responsiveness to PPH emergencies.

## II. MATERIALS AND METHODS

### Design and participants

This was a quantitative 1:2 unmatched case–control study conducted across six public hospitals in Lusaka, Zambia that is, five first-level hospitals and one tertiary referral facility. We enrolled 318 women (106 cases and 212 controls). Cases met the WHO definition of primary PPH as estimated blood loss ≥ 500 ml within 24 hours after vaginal delivery. Controls were contemporaneous vaginal deliveries without PPH. Exclusion criteria included conditions affecting coagulation or predisposition to bleeding, such as sickle cell disease, hemophilia, severe pre-existing anemia, or major cardiac disease. Participants were selected using systematic sampling, choosing every second eligible woman until facility quotas were filled. Proportional sampling ensured representation across the six sites

### Procedure and Instruments

After written informed consent, trained data collectors administered a pre-tested questionnaire and abstracted clinical data from medical records using a standardized form. Blood loss estimation was applied uniformly to all participants. Visual assessment was supplemented by gravimetric measurement (weighing soaked materials and converting weights to milliliters using a standard conversion) and, where applicable, by measuring blood collected through a suction machine. This ensured consistency in the definition of primary postpartum hemorrhage across cases and controls. Genital tract trauma was classified from clinical inspection and delivery notes, recording cervical tears, perineal tears by standard grading, and vaginal lacerations. An obstetrics and gynecology consultant conducted a central orientation for research assistants to improve measurement reliability. Data collection procedures and tools were uniform across sites following a central training workshop.

### Data Analysis

Data were coded and entered into Statistical Package for the Social Sciences (SPSS) version 25 for analysis. Descriptive statistics (frequencies, percentages, means, and standard deviations) were used to summarize participant characteristics and maternal outcomes. Inferential statistics included correlation analysis to examine relationships between accessibility, provider competence, and outcomes, and regression analysis to identify predictors of maternal outcomes. Statistical significance was set at *p* < 0.05.

### Ethical Considerations

Ethical clearance for the study was obtained from the University of Zambia Biomedical Research Ethics Committee (UNZABREC) (REF: no 074-2019). Additional permission was secured from the National Health Research Authority (NHRA) and the Ministry of Health, Zambia. Written informed consent was obtained from all participants prior to data collection. Confidentiality and anonymity were maintained throughout the study, with data stored securely and used solely for research purposes.

## III. RESULTS

**Table 1** summarizes the sociodemographic characteristics of study participants. Cases (N = 106) and controls (N = 212) were comparable across all measured baseline variables. Median age was similar between groups (27 years [IQR: 22–32] for cases vs. 27 years [IQR: 22.5–32] for controls; p = 0.636). No statistically significant differences were observed in education level (p = 0.658), employment status (p = 0.870), residence category (p = 0.158), marital status (p = 0.683), or HIV status (p = 0.516), indicating a well-balanced study population suitable for comparative analysis.

**Table 1.**
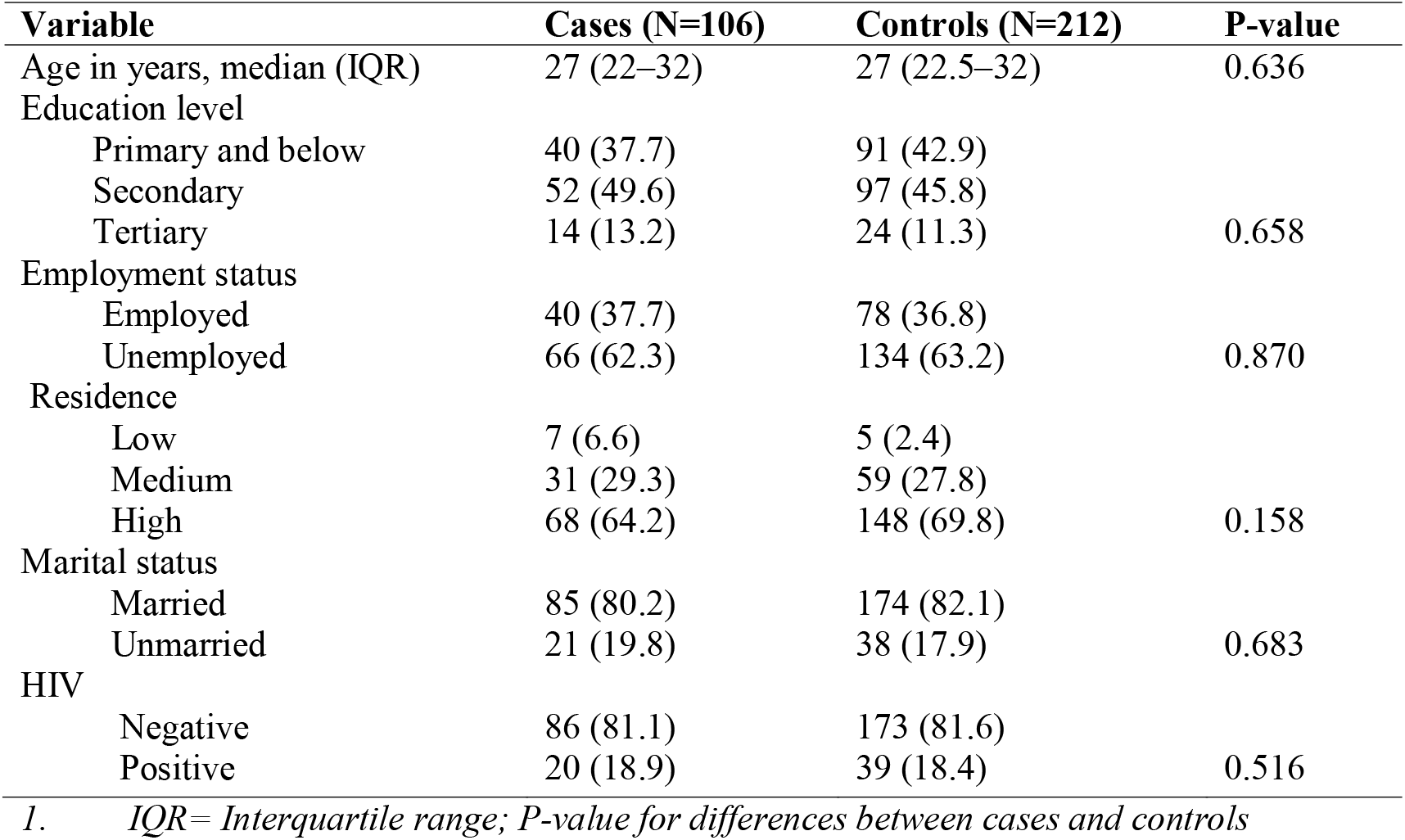
Participants’ characteristics.

**Table 2** highlights that women who experienced primary postpartum hemorrhage (PPH) had significantly worse outcomes compared to controls across all measured indicators. Among the 106 cases, 56.8% (n=60) required blood transfusion, while none of the 212 controls did, highlighting the direct link between hemorrhage and blood product demand. Hemorrhagic shock occurred in 11% (n=12) of cases, hysterectomy in 3.8% (n=4), and maternal mortality in 3.8% (n=4), all absent in controls. The high rates of referral 29.2%, (n=31), SOU admission 10.4%, (n=11), and ICU admission 5.7%, (n=6) among cases underscore the systemic burden PPH places on emergency obstetric care and the urgent need for improved preparedness at first-level hospitals.

**Table 2.** Descriptive Statistics of Maternal Outcomes Following Primary PPH.

| Outcome Variable | Cases (N=106) | Controls (N=212) | P-value |
| --- | --- | --- | --- |
| Blood transfusion (%) | 56.8 | 0.0 | <0.001 |
| Hemorrhagic shock (%) | 11.0 | 0.0 | <0.001 |
| Hysterectomy (%) | 3.8 | 0.0 | 0.045 |
| Referral to tertiary (%) | 29.2 | 0.0 | <0.001 |
| SOU admission (%) | 10.4 | 0.0 | <0.001 |
| ICU admission (%) | 5.7 | 0.0 | 0.002 |
| Maternal mortality (%) | 3.8 | 0.0 | 0.021 |

*N= Number of subjects*

**Table 3** presents the correlation analysis which revealed that both accessibility of care and provider competence were negatively associated with adverse maternal outcomes. Facilities with stronger infrastructure and better-trained staff reported fewer complications, with accessibility showing a significant negative correlation with transfusion (r = -0.42, p < 0.01) and referral (r = - 0.48, p < 0.01). Provider competence also correlated negatively with mortality (r = -0.33, p < 0.05), suggesting that improved training reduces fatal outcomes. These findings indicate that strengthening lower-level hospitals and investing in continuous training for providers could substantially reduce the severity of PPH outcomes, thereby improving maternal survival rates.

**Table 3.**
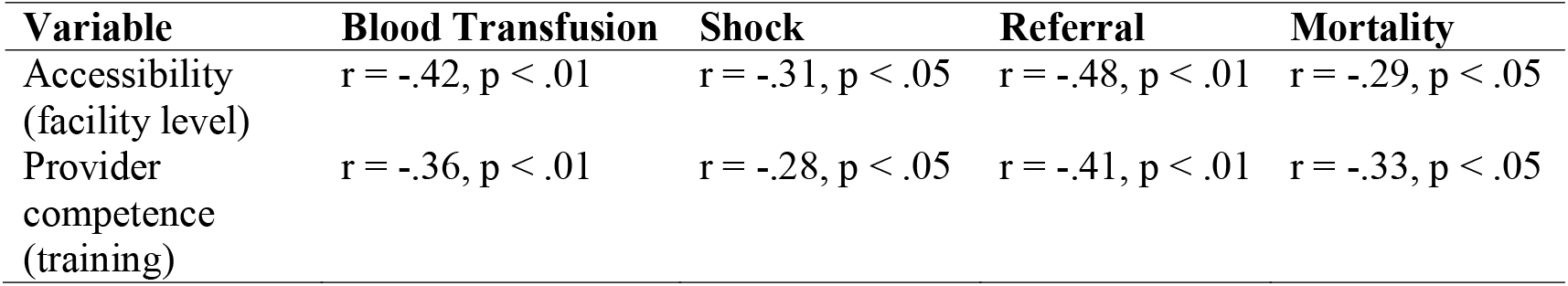
Correlation Between Accessibility, Provider Competence, and Maternal Outcomes.

Regression analysis in **table 4** identified delays in referral and lack of blood availability as the strongest predictors of maternal mortality. Women with uterine atony were nearly three times more likely to die (OR = 2.8, 95% CI: 1.4–5.6, p = 0.003), while those referred after more than two hours had a 3.5-fold increased risk (95% CI: 1.7–7.2, p < 0.001). Lack of blood availability was the most critical factor, with an odds ratio of 4.2 (95% CI: 2.0–8.9, p < 0.001). These results emphasize that timely referral systems and reliable blood transfusion services are critical determinants of survival, and addressing these systemic gaps could significantly reduce maternal deaths from PPH.

**Table 4.** Logistic Regression Predicting Maternal Mortality.

| Predictor Variable | Odds Ratio (OR) | 95% CI | P-value |
| --- | --- | --- | --- |
| Uterine atony | 2.8 | 1.4 – 5.6 | 0.003 |
| Cervical/vaginal tears | 1.6 | 0.9 – 3.1 | 0.087 |
| Delay in referral (>2 hrs) | 3.5 | 1.7 – 7.2 | <0.001 |
| Lack of blood availability | 4.2 | 2.0 – 8.9 | <0.001 |

The comparative **table 5** highlights stark differences between PPH cases and controls, with adverse outcomes occurring exclusively among cases. Blood transfusion was required in 56.8% (n=60) of cases, referral in 29.2% (n=31), and SOU/ICU admission in 16.1% (n=17 combined), while none of the controls experienced these complications. Maternal mortality was observed in 3.8% (n=4) of cases, compared to 0% in controls. This clear disparity illustrates the disproportionate burden PPH places on maternal health systems and reinforces the need for proactive strategies to prevent hemorrhage and strengthen emergency response capacity.

**Table 5.**
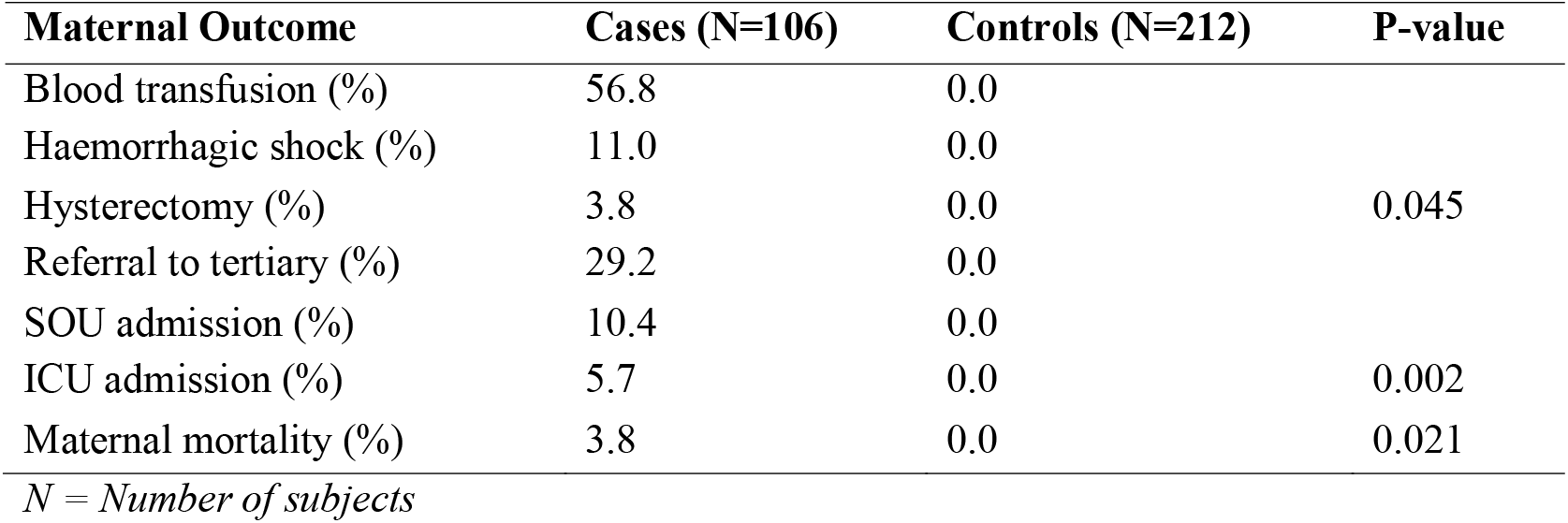
Comparative Maternal Outcomes in PPH Cases vs. Controls.

## IV. DISCUSSION

The outcomes of postpartum hemorrhage (PPH) examined in this study included the need for blood transfusion, hemorrhagic shock, hysterectomy, referral, admission to the special observation unit (SOU) or intensive care unit (ICU), and maternal mortality. While cervical tears were identified as the most frequent immediate cause of primary PPH, uterine atony accounted for the majority of secondary complications, consistent with global evidence that uterine atony remains the leading contributor to severe maternal morbidity.^[20]^ Several studies have highlighted the obstetric shock index (SI) as a more reliable predictor of adverse outcomes compared to traditional vital signs, facilitating earlier resuscitation and improved survival.^[21,22]^ Recent prospective analyses further support SI’s predictive utility in low-resource settings, though some findings caution that its sensitivity may vary depending on population characteristics and comorbidities.^[23,24]^ Taken together, these findings underscore both the promise and limitations of SI as a clinical tool, while reaffirming uterine atony as the predominant driver of poor outcomes in PPH.

### Blood transfusion and primary PPH

The need for blood transfusion remains one of the most significant complications of primary PPH, directly influencing maternal outcomes. In this study, transfusion was required in 56.8% of cases, a figure comparable to findings from Mali, where 42.6% of women with PPH required transfusion.^[25,26]^ More recent evidence from sub-Saharan Africa continues to highlight high transfusion rates, reflecting persistent challenges in timely access to blood products.^[11]^ In contrast, data from high-income countries demonstrate substantially lower transfusion requirements, with a nationwide study in the Netherlands reporting rates between 4.1% and 6.4%.^[27]^ Similarly, a Norwegian cohort study found that improved antenatal care and active management of the third stage of labor significantly reduced transfusion needs.^[20]^

The discrepancy between low- and high-resource settings may be explained by differences in antenatal care coverage, blood bank infrastructure, and predictors of PPH severity. Severe PPH consistently emerges as a strong predictor of transfusion, with recent systematic reviews reaffirming this association.^[28]^ However, some studies caution that transfusion practices vary widely depending on clinical thresholds and local guidelines, which may lead to under- or over- utilization of blood products.^[24]^ Taken together, these findings suggest that while transfusion remains a critical intervention in managing PPH, its frequency is shaped by both health system capacity and clinical decision-making, underscoring the need for context-specific strategies to optimize maternal outcomes.

### Hemorrhagic shock and primary PPH

Following primary PPH, women may develop hemorrhagic shock characterized by inadequate tissue perfusion and insufficient oxygen delivery to vital organs.^[29]^ In this study, 11% of cases experienced hemorrhagic shock, while none occurred among controls. Recent evidence underscores that the severity of shock is closely linked to the volume of blood loss, with mild shock typically occurring after a 20% reduction in circulating blood volume, leading to early compromise of organ perfusion.^[30,31]^

Successful management of hemorrhagic shock has increasingly been associated with multidisciplinary collaboration and rapid initiation of resuscitation protocols. Updated Canadian guidelines emphasize team-based approaches, including early fluid resuscitation, blood product administration, and monitoring guided by clinical endpoints such as urine output and pulse pressure.^[32]^ Similarly, systematic reviews highlight that outcomes improve significantly when shock is recognized early and managed with standardized bundles of care.^[28]^ However, some studies caution that despite advances, variability in recognition and intervention thresholds across health systems continue to contribute to preventable morbidity and mortality.^[33]^

### Hysterectomy and primary PPH

We observed hysterectomy in 3.8% of cases, though the small sample size limited statistical association with PPH. Recent hospital-based studies in Europe report lower rates of peripartum hysterectomy, with Norway documenting incidences below 1% due to improved preventive measures such as uterotonics, balloon tamponade, and multidisciplinary emergency protocols.^[20]^ Similarly, updated reviews from high-resource settings confirm a continued decline in hysterectomy rates, reflecting advances in conservative surgical techniques and standardized bundles of care.^[34]^

By contrast, findings from low- and middle-income countries (LMICs) highlight substantially higher hysterectomy rates. A retrospective study in Tunisia reported rates exceeding 3% in secondary hospitals, largely attributed to delays in accessing emergency care and limited blood product availability.^[35]^ African systematic reviews also emphasize that peripartum hysterectomy remains a critical life-saving intervention where health system capacity is constrained, with maternal morbidity and mortality disproportionately elevated.^[33]^

### Need for referral

This study showed that PPH was responsible for a considerable number of referrals requiring advanced management, including blood transfusions, surgical interventions such as hysterectomy, and intensive care for critically ill patients. Among women with primary PPH, 29.2% were referred for further management to the WNH. This finding aligns with evidence from India, which reported referral rates of 32.1% among women with severe PPH, underscoring the burden on tertiary facilities. Most referrals following PPH are often linked to inadequate management at peripheral health centers or incomplete prenatal care, as highlighted in a Madagascan study.^[36]^

Recent African data further support these observations, showing that delays in referral and gaps in emergency transport systems significantly contribute to adverse maternal outcomes.^[33]^ Similarly, a Zambian study emphasized that strengthening referral pathways and ensuring timely access to blood products are critical to reducing preventable morbidity and mortality.^[11]^ In addition, systematic reviews of obstetric critical care admissions confirm that referral patterns are strongly associated with the severity of hemorrhage and the availability of specialized interventions at higher-level facilities.^[28]^

### Admission to special observation unit/ intensive care unit and primary PPH

In this study, 10.4% of participants were admitted to the special observation unit (SOU) and 5.7% to the intensive care unit (ICU). The SOU serves as an intermediate level of care where critically ill obstetric patients are closely monitored before transfer to ICU or while awaiting ICU availability. Primary postpartum hemorrhage (PPH) is a well-recognized cause of SOU and ICU admission following massive bleeding. Recent cohort studies confirm that severe PPH remains one of the leading indications for obstetric critical care admissions globally, with ICU utilization rates varying widely depending on health system capacity.^[28]^

Evidence from China and other Asian settings highlights increasing ICU admissions due to PPH, particularly in severe cases where delayed recognition or inadequate initial management exacerbates outcomes.^[37,38]^ Similarly, systematic reviews from Africa demonstrate that women referred to ICU following PPH are often already in shock on arrival, reflecting gaps in referral pathways and emergency transport systems.^[33]^ The discrepancy in admission proportions between this study and others may be explained by differences in admission guidelines, availability of ICU beds, and thresholds for escalation of care.

Successful intervention measures reported in recent guidelines emphasize immediate resuscitation directed by clinical endpoints such as urine output, pulse rate, and blood pressure, alongside early blood transfusion and surgical interventions when indicated.^[32]^ Collectively, these findings underscore the critical role of SOU and ICU in managing severe PPH, while highlighting the need for standardized admission criteria and strengthened referral systems to reduce preventable maternal morbidity and mortality.

### Maternal mortality

Maternal mortality remains a preventable yet persistent public health challenge, with postpartum hemorrhage (PPH) continuing to be one of the leading causes of death worldwide. In this study, maternal mortality following primary PPH was 3.8%. Recent systematic reviews across sub-Saharan Africa confirm that PPH accounts for a substantial proportion of maternal deaths, often exceeding 20% in severe cases, particularly where delays in referral and limited access to blood products are common.^[39,33]^

Global burden analyses further highlight stark disparities: while high-income countries report markedly reduced mortality due to standardized bundles of care and improved prenatal coverage, low- and middle-income countries continue to experience disproportionately high rates.^[40]^ Evidence from Mozambique demonstrates that multi-faceted interventions, including improved detection and rapid escalation of care, significantly reduce mortality, underscoring the importance of system-level strengthening.^[41]^

The discrepancy in mortality rates between this study and others may be explained by differences in health system capacity, study duration, and access to emergency obstetric care. Updated WHO recommendations emphasize earlier detection, bundled management strategies, and equitable access to uterotonics and blood products as critical steps toward reducing preventable maternal deaths.^[42]^ Collectively, these findings reaffirm that while maternal mortality from PPH is largely preventable, achieving reductions requires both clinical innovation and systemic improvements in referral pathways and resource allocation.

## V. CONCLUSION

Postpartum hemorrhage (PPH) following vaginal delivery was strongly associated with severe maternal outcomes, including high rates of blood transfusion, hemorrhagic shock, referral to tertiary care, and maternal mortality. These complications occurred exclusively among PPH cases, underscoring the disproportionate burden placed on first-level hospitals with limited emergency capacity. Regression analysis identified delays in referral and lack of blood availability as the most critical predictors of mortality, while provider competence and accessibility of care emerged as protective factors. These findings highlight the urgent need for system-level interventions to strengthen blood transfusion services, streamline referral pathways, and ensure adherence to standardized national guidelines for PPH management. Continuous training of midwives and frontline staff, investment in emergency preparedness, and integration of monitoring tools are essential to improving maternal survival. Future research should employ longitudinal designs and larger, more diverse samples across different levels of care to validate these findings. Comparative studies across regions, alongside evaluations of cost-effective health system interventions such as blood bank strengthening and referral optimization, will be critical to reducing preventable maternal deaths. Ultimately, addressing the systemic gaps identified in this study offers a pathway toward safer childbirth and improved maternal health outcomes in resource-constrained settings.

## Data Availability

All data produced in the present study are available upon reasonable request to the authors

## Acknowledgements

The authors gratefully acknowledge the substantive contributions of the midwives and medical officers from the participating hospitals in Lusaka, Zambia, who assisted with data collection and patient care during the study. Their support was provided as part of their professional duties at the Lusaka District Health Office and the Women and Newborn Hospital, without additional financial compensation or external funding.

